# Acquisition Speed versus Spatial Resolution in Ultra-High-Resolution Photon-Counting CT: Phantom Study to Guide Protocol Development

**DOI:** 10.64898/2026.10.04.26363713

**Authors:** Navid Azimi, Joao A.C. Lima, Amir Pourmorteza

## Abstract

**Background:** Faster ultrahigh-resolution (UHR) photon-counting CT acquisitions facilitate breath-hold imaging, but identical reconstruction settings may conceal differences in spatial resolution.

**Purpose:** To assess spatial resolution and depiction of small airways and venous stents across helical UHR acquisition configurations.

**Materials and Methods:** Stationary wire, airway, and venous-stent phantoms underwent one acquisition per configuration using five configurations on a dual-source scanner at 120 kVp. Reconstructions used 0.2- and 0.4-mm thicknesses, the same sharp kernel, and iterative reconstruction strength 1. We measured the full-width at half-maximum of the slice sensitivity profile (SSP) and spatial frequency at 20% of the modulation transfer function (MTF20). Rank-based F tests with Holm adjustment assessed configuration differences in resolution and variability across slices. Airway detection used a profile contrast-to-noise criterion; stent assessment was qualitative.

**Results:** Calculated acquisition time for 400-mm coverage decreased from 9.80 to 1.30 seconds between the slowest and fastest configurations. At 0.2-mm reconstruction thickness, median SSP increased from 0.316 to 0.496 mm, and median MTF20 decreased from 2.870 to 2.059 lp/mm (both P<.001). Interquartile ranges increased from 0.047 to 0.197 mm for SSP and from 0.256 to 0.654 lp/mm for MTF20 (both variability P<.001). Increasing reconstruction thickness to 0.4 mm increased SSP by a median 0.221 mm across 2459 matched slice-wire measurements (P<.001). At 0.2-mm reconstruction thickness, 0.4- and 0.6-mm lumens with 0.2-mm walls were detected only in the slowest configuration. Faster configurations showed less distinct stent struts and greater adjacent-wall distortion.

**Conclusion:** Faster helical UHR photon-counting CT configurations had poorer spatial resolution despite identical nominal slice thickness. Consistent acquisition and reconstruction settings are important for longitudinal comparisons.

**Original Research:** *Summary Statement:* Faster helical ultrahigh-resolution photon-counting CT configurations had poorer spatial resolution and small-structure visibility despite identical reconstruction settings, supporting task-specific protocols and consistent techniques for longitudinal imaging.

*Key Results:*

1. At 0.2-mm reconstruction thickness in this phantom study, median slice sensitivity profile was 0.496 versus 0.316 mm for the fastest versus slowest configurations (calculated 400-mm scan times, 1.30 vs 9.80 seconds; P<.001).
2. In-plane spatial frequency at 20% modulation decreased from 2.870 to 2.059 lp/mm (P<.001), with greater variability across slices.
3. At 0.2-mm reconstruction thickness, only the slowest configuration depicted 0.4- and 0.6-mm lumens with 0.2-mm walls.

## INTRODUCTION

Photon-counting detector CT (PCCT) promises finer spatial resolution, spectral information, lower image noise, and improved dose efficiency (1). Early studies confirmed gains in fine-structure visualization and quantitative imaging with ultrahigh-resolution (UHR) PCCT compared with energy-integrating detector CT (2, 3). In practice, however, these advantages can compete. PCCT has also been implemented at higher rather than lower radiation dose in clinical practice (4). Its principal advantages cannot necessarily be maximized simultaneously within a single acquisition(5).

Current commercial cadmium telluride (CdTe) PCCT scanners cover less anatomy per rotation in UHR mode than in standard-resolution mode. For chest and abdomen examinations, this narrower coverage must be balanced against completing the scan within a breath-hold. Increasing acquisition speed helps meet this requirement but can degrade the spatial resolution that motivated UHR imaging (6, 7). A nominal 0.2-mm slice thickness or UHR designation does not ensure that the corresponding spatial resolution is achieved. Images reconstructed with the same slice thickness and kernel may differ in their ability to depict small structures. This distinction matters in longitudinal studies, where differences in technique could complicate assessment of disease progression or treatment response. Consistent acquisition and reconstruction settings are therefore important when comparing examinations over time.

The purpose of this phantom study was to measure in-plane and through-plane spatial resolution across clinically available UHR acquisition configurations and relate these measurements to scan time and depiction of small airways and vascular stents. Our aim was to inform UHR chest and abdomen protocol development, including selection of consistent techniques for longitudinal studies.

## MATERIALS AND METHODS

### Study Design and Test Objects

We tested a dual-source cadmium-telluride PCCT system (NAEOTOM Alpha Peak; Siemens Healthineers, Forchheim, Germany), with Syngo CT VB20A software, at Emory University. Each phantom underwent one acquisition per configuration.

A Revolution QA phantom (model 137856101, part no. 5477995, Rev 8; GE HealthCare, Waukesha, WI) contained five 0.05-mm tungsten wires: four tilted at a 1:2 slope (26.57° relative to z) and one parallel to z. The tilted wires were arranged on two horizontal lines 6 cm above and below isocenter. The parallel wire was 3.5 cm from isocenter. We corrected for a 6° residual phantom roll.

We designed a separate 8×7 airway matrix in SolidWorks (Dassault Systèmes SolidWorks Corporation, Waltham, MA), with lumen diameters of 0.1–3.0 mm and wall thicknesses of 0.1–1.5 mm. We printed it by stereolithography (Mono 4K; Anycubic, Shenzhen, China) using clear green resin (ABS-Like Resin Pro 2; Anycubic). The matrix was oblique to x, y, and z, with the lumen-diameter direction more closely aligned with z.

Three venous stents were embedded in 20% synthetic ballistic gel (Clear Ballistics, Greenville, SC) within a nominal 3-inch polyvinylchloride pipe. The main comparison used a 10×60-mm nitinol venous stent (Venovo; Bard/BD, Tempe, AZ); two additional venous stents are shown in the Supplement. We placed the pipe in the insert opening of the Dynamic Cardiac Phantom (Model 008C; CIRS, Norfolk, VA). The same phantom held the airway matrix for separate scans. All objects were within 10 cm of isocenter and were scanned without cardiac or respiratory motion.

### Scan and Reconstruction Parameters

We tested five helical UHR configurations (C1–C5) at 120 kVp. Details are provided in Table 1. For C1, volume CT dose index (CTDIvol) was 6.45 mGy for the wire phantom and 6.41 mGy for the stent and airway phantoms, a 0.6% difference.

**Table 1.** Acquisition parameters.

| Parameter | C1 | C2 | C3 | C4 | C5 |
| --- | --- | --- | --- | --- | --- |
| UHR acquisition mode | SS UHR | Flash UHR | Flash UHR | Flash UHR | Flash Wide UHR |
| Source mode | Single source | Dual source | Dual source | Dual source | Dual source |
| Detector collimation [mm] | $120 \times 0.2$<br>(24.0) | $96 \times 0.2$<br>(19.2) | $96 \times 0.2$<br>(19.2) | $96 \times 0.2$<br>(19.2) | $120 \times 0.2$<br>(24.0) |
| Helical pitch | 0.85 | 1.00 | 2.00 | 3.20 | 3.20 |
| Rotation time [s] | 0.50 | 0.25 | 0.25 | 0.25 | 0.25 |
| Table speed [mm/s] | 40.8 | 76.8 | 153.6 | 245.6 | 307.2 |
| Calculated acquisition time for 400-mm coverage [s] | 9.80 | 5.21 | 2.60 | 1.63 | 1.30 |
| Tube voltage [kV] | 120 | 120 | 120 | 120 | 120 |
| Focal spot (class: dimensions) [mm] | XS: $0.4 \times 0.4$ | XS: $0.4 \times 0.4$ | S: $0.6 \times 0.7$ | M: $0.8 \times 1.1$ | M: $0.8 \times 1.1$ |
| Total tube current [mA] | 136 | 320 | 639 | 1023 | 1055 |
| Source A/Source B current [mA] | 136 | 160/160 | 320/320 | 511/511 | 527/527 |
| Total effective exposure [mAs] | 80 | 80 | 80 | 80 | 82 |
| Source A/Source B effective exposure [mAs] | 80 | 40/40 | 40/40 | 40/40 | 41/41 |
| CTDIvol [mGy] | 6.413 | 6.558 | 6.559 | 6.558 | 6.612 |

Parameters are from the airway phantom acquisitions.
Tube current, effective mAs, and CTDIvol are representative displayed values because longitudinal tube-current modulation was enabled.
Total and source-specific tube current and effective mAs are retained as recorded. Source-specific values are reported as A/B; single-source values are reported once. Reported total and source-specific currents may not sum exactly.
UHR = ultra-high resolution.
Acquisition time for 400-mm coverage was calculated using table speed reported in the DICOM header.

Images were reconstructed from all detected x-ray photons above the low-energy threshold (T3D) at 0.2- and 0.4-mm slice thickness with matching increments, a 1024×1024 matrix, Qr89u (the sharpest quantitative kernel available), and quantum iterative reconstruction (QIR) strength 1. Reconstruction field of view (FOV) was 151.8 mm for wires and 170 mm for airways and stents, with in-plane voxel sizes of 0.148×0.148 and 0.166×0.166 mm², respectively, small enough to limit partial-volume effects from image sampling. We also assessed sensitivity to QIR strength using 0.4-mm reconstructions.

### Quantitative Image Analysis

We analyzed images in MATLAB R2026a (MathWorks, Natick, MA). Through-plane resolution was defined as the full-width at half-maximum (FWHM) of the slice sensitivity profile (SSP), measured using the multiple tilted-wire method (8). Orthogonal Gaussian fits were corrected for wire tilt and phantom roll. Larger SSP values indicate poorer through-plane resolution (Fig 1A).

**Figure 1.**
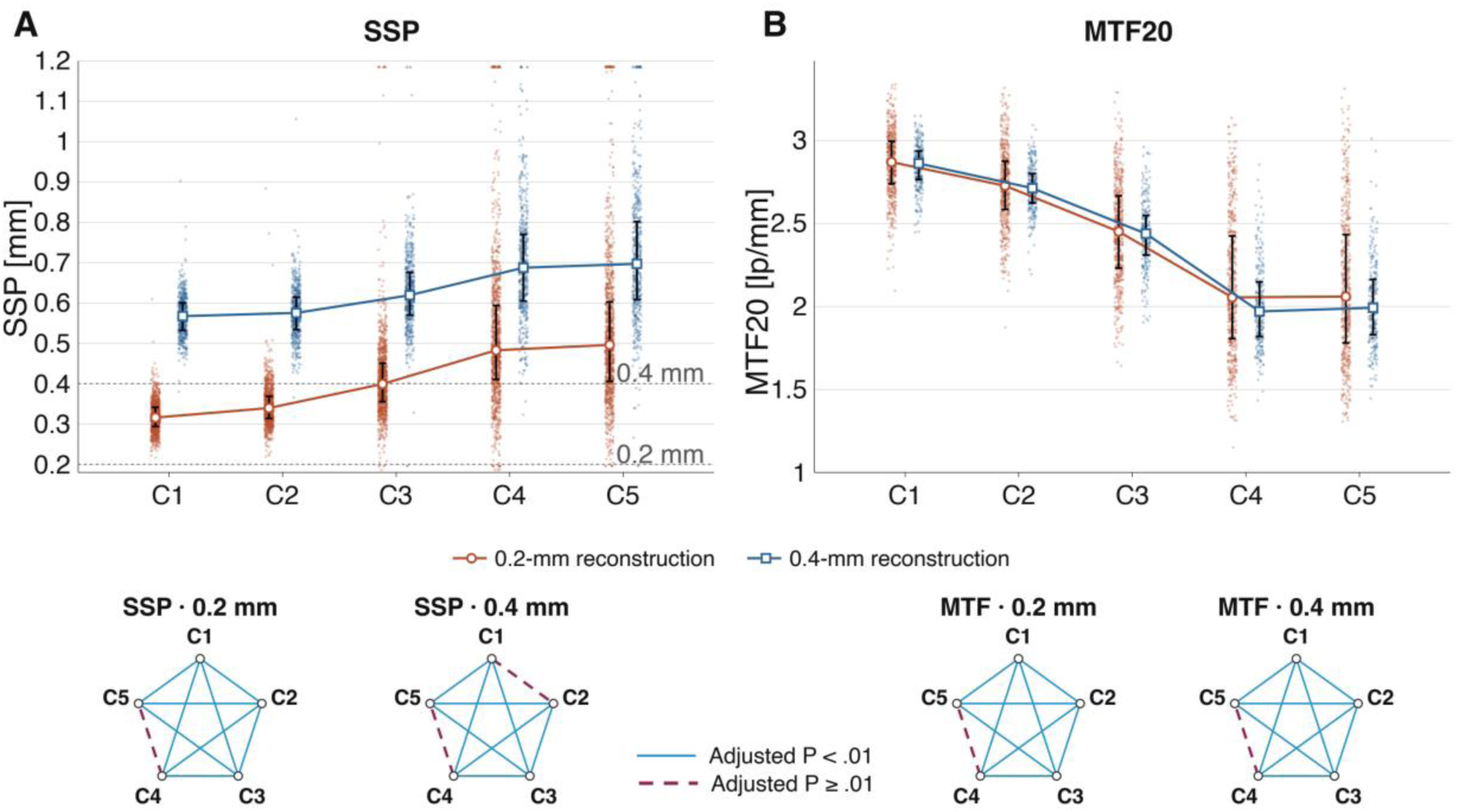
Through-plane and in-plane spatial resolution across acquisition configurations. SSP (A) and slice-level MTF20 (B) are shown for 0.2- and 0.4-mm reconstructions using QIR 1. Points show individual measurements; colored lines and markers show medians, and black error bars show IQRs. Dotted horizontal lines in A indicate 0.2 and 0.4 mm. Triangles at axis limits indicate retained observations outside the displayed range. Pentagram edges summarize pairwise configuration comparisons for each endpoint and reconstruction thickness: solid blue lines indicate Holm-adjusted P<.01 and dashed burgundy lines indicate P≥.01. Complete MTF50 and MTF20 results are shown in Figure S2. IQR = interquartile range; MTF = modulation transfer function; QIR = quantum iterative reconstruction; SSP = slice sensitivity profile.

We measured the in-plane modulation transfer function (MTF) from the parallel wire (9). The background-corrected two-dimensional wire response in each slice was Fourier transformed and radially averaged, with noise-power correction and a monotonic constraint. MTF50 and MTF20 were the frequencies at 50% and 20% of the zero-frequency response. Mean curves were calculated for display. Lower MTF50 and MTF20 values indicate poorer in-plane resolution; MTF20 is shown in Figure 1B.

To illustrate how in-plane and through-plane resolution jointly affect a submillimeter object’s appearance, we simulated a 0.5-mm-diameter circle in the x-z plane (Fig 2). Each configuration’s mean MTF specified x-direction blurring; a Gaussian response with FWHM equal to its median SSP specified z-direction blurring. All images used a common grayscale normalized to ideal-object contrast. The simulation illustrated blurring without modeling noise or helical distortion.

**Figure 2.**
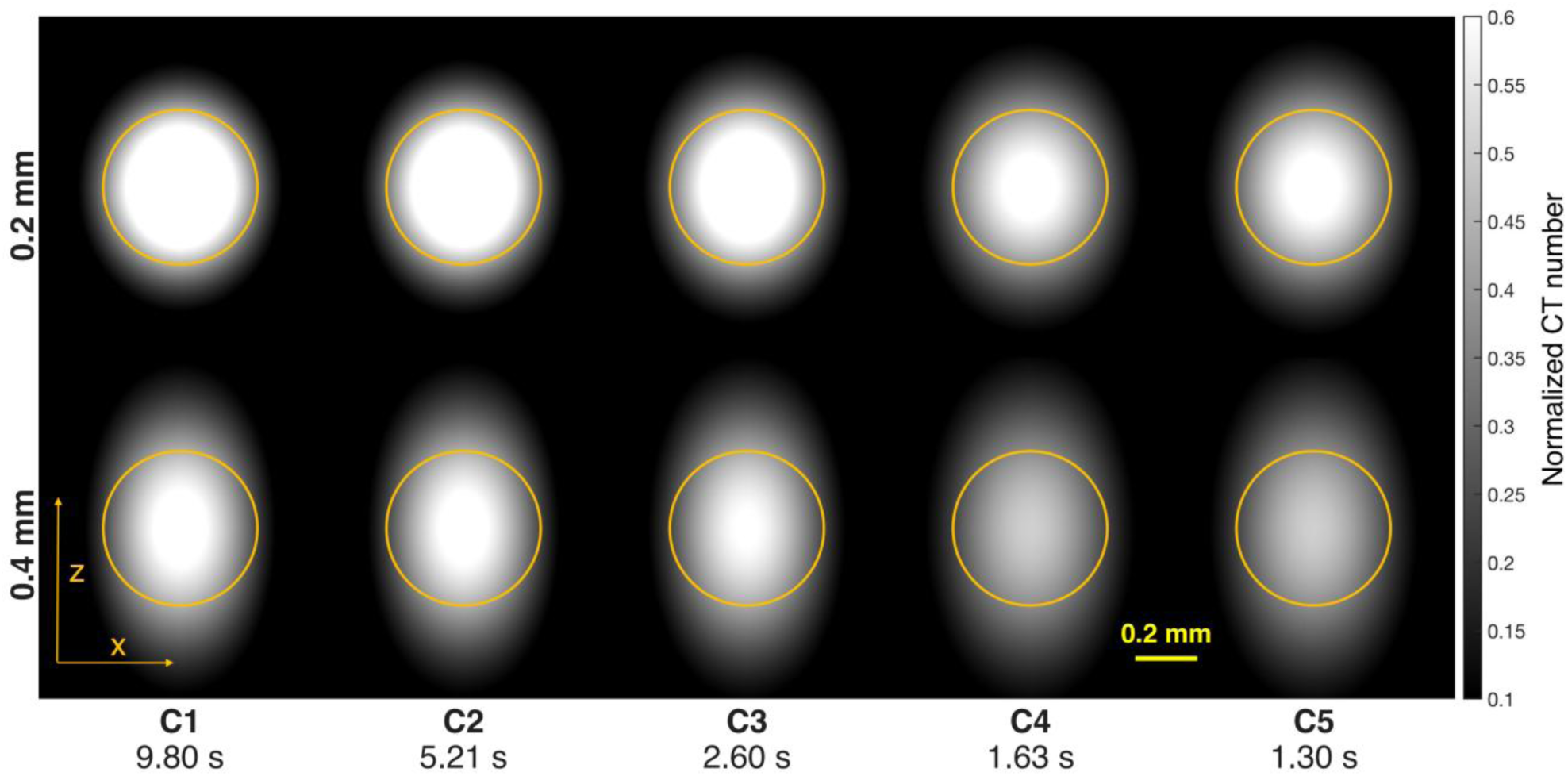
Combined effects of in-plane and through-plane spatial resolution. A simulated 0.5-mm-diameter circular object was blurred along x using each configuration’s measured mean MTF magnitude and along z using a Gaussian response with full width at half maximum equal to the measured median SSP. Yellow circles mark the ideal-object boundary. Results are shown for 0.2-mm (top) and 0.4-mm (bottom) reconstructions using QIR 1. Columns show C1–C5. Labels report calculated time for 400-mm coverage. The common grayscale is normalized to ideal-object contrast. Yellow arrows identify x and z; scale bar, 0.2 mm. The simulation illustrates anisotropic blurring and does not model noise or helical distortion. MTF = modulation transfer function; QIR = quantum iterative reconstruction; SSP = slice sensitivity profile.

Airway detection was assessed on matched 5-mm oblique mean-intensity reformations, with lumen diameter and wall thickness identified in the matrix schematic (Fig 3). We sampled two orthogonal profiles through each airway center. Contrast was the mean of the two wall peaks minus the lumen minimum. Detection required mean contrast across the profiles to exceed four times the mean surrounding-air standard deviation. We selected the contrast-to-noise ratio threshold of 4 within the commonly cited Rose range of 3–5 (10). We mapped detection by lumen diameter and wall thickness. Damaged or unevaluable positions were excluded from analysis and CT panels but were retained in the schematic.

**Figure 3.**
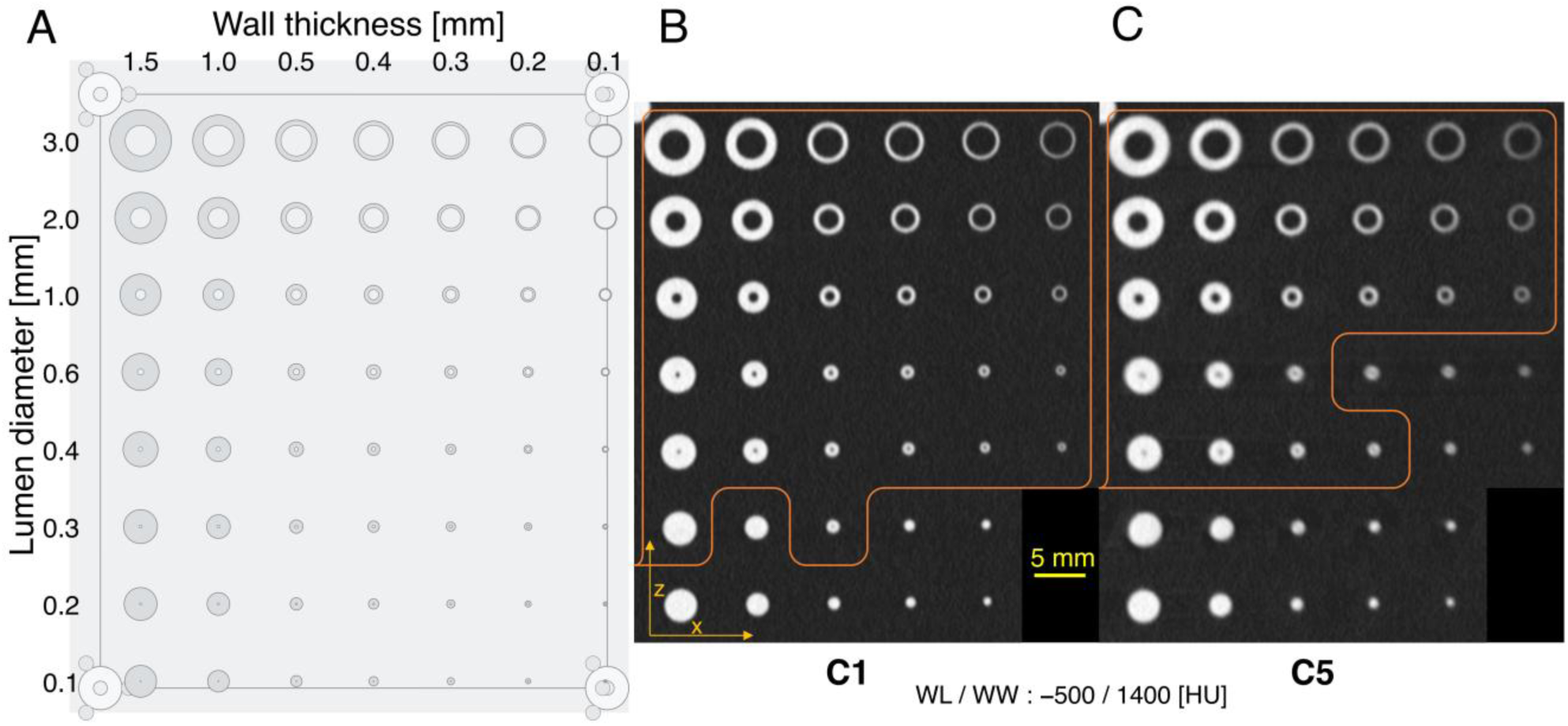
Airway-matrix design and lumen conspicuity in C1 and C5. The schematic (left) shows nominal lumen diameters by row and wall thicknesses by column. Matched 5-mm oblique mean-intensity reformations from 0.2-mm reconstructions show C1 (center) and C5 (right) at equal magnification. Orange contours mark lumen detectability boundaries at the sampled positions. They are visual guides, retain isolated nonmonotonic findings, and do not represent fitted continuous thresholds. Damaged or unevaluable positions are omitted from CT panels and retained in the schematic. Window level/width, −500/1400 HU; scale bar, 5 mm.

Lumen detectability boundaries were visual guides through the sampled lumen-diameter and wall-thickness combinations, retained isolated nonmonotonic findings, and were not fitted continuous thresholds. Excluded positions included the damaged 0.1-mm-wall column and lowest lumen row. Unclassified positions remained unclassified; we did not report pooled detection proportions or inferential comparisons.

### Qualitative Image Analysis

At matched display settings, we compared airway cross-sections and longitudinal reformations (Figs 4, 5), axial stent images, and longitudinal stent maximum-intensity projections (MIPs). For the main stent, matched 7-mm MIPs showed strut definition and the geometry of the adjacent phantom pipe wall (Fig 6).

**Figure 4.**
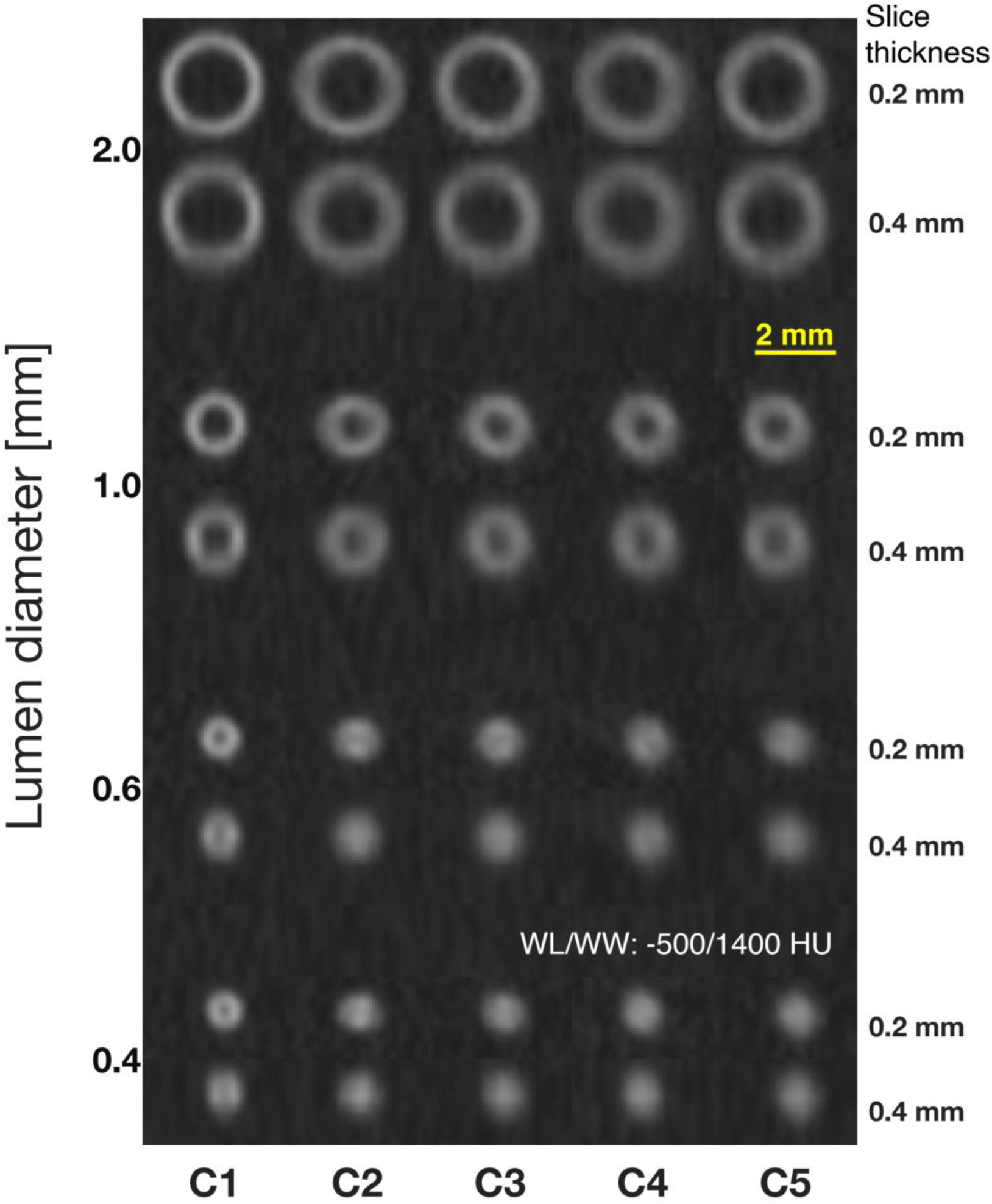
Airway detail at two reconstruction thicknesses. Cross-sectional enlargements show airways with 0.2-mm walls and nominal lumen diameters of 2.0, 1.0, 0.6, and 0.4 mm. C1–C5 run from left to right. For each lumen diameter, the 0.2-mm reconstruction is above the corresponding 0.4-mm reconstruction. The smallest lumens lose conspicuity in the faster configurations. Images share magnification and window level/width of −500/1400 HU; scale bar, 2 mm.

### Statistical Analysis

We summarized SSP, MTF50, and MTF20 as median (interquartile range [IQR]) and compared resolution and variability across C1–C5 separately at each slice thickness. The 0.2-mm reconstructions were the primary comparison, with 0.4-mm reconstructions assessed secondarily. Analyses used slice-wire measurements for SSP and individual slices for MTF.

We tested configuration differences using F tests on rank-normal scores, including physical wire as a fixed blocking factor for SSP. To compare variability, we tested rank-normalized absolute deviations from group medians, with within-wire centering and wire adjustment for SSP, adapting median-centered variance testing (11). Pairwise configuration comparisons and variability comparisons with C1 underwent separate Holm adjustments for each endpoint and slice thickness (12). A paired Wilcoxon test compared SSP between slice thicknesses. Tests were two-sided, with P<.01 considered statistically significant.

QIR comparisons used strengths 0–2 (0–1 for C1) and measurements valid at every available strength. SSP measurements were matched by wire and z position, and MTF by slice index. Friedman tests compared QIR levels; C1 used a paired Wilcoxon signed-rank test. Repeated-measures Levene tests assessed absolute deviations from QIR-group medians, with separate centering by wire for SSP and Greenhouse–Geisser correction. C1 used a paired median-centered Levene test. Levene testing has been used to compare variability in CT measurements (13). P values were Holm-adjusted across configurations separately for each endpoint and outcome. IQRs described variability; Levene tests did not test equality of IQRs.

## RESULTS

### Acquisition Speed and Spatial Resolution

Calculated acquisition time for a 400-mm scan decreased from 9.80 seconds in C1 to 1.30 seconds in C5 (Table 1). At 0.2-mm slice thickness, median SSP increased from 0.316 (0.294–0.341) mm to 0.496 (0.405–0.602) mm, a 57.0% increase (Table 2, Fig 1A). Median MTF20 decreased from 2.870 (2.739–2.995) lp/mm to 2.059 (1.780–2.434) lp/mm, a 28.2% decrease (Fig 1B). All pairwise comparisons had P<.001 except C4 versus C5 (SSP, P=.460; MTF20, P=.983).

**Table 2.** SSP and slice-level MTF20 across acquisition configurations.

| Config. | Thickness [mm] | SSP n | SSP [mm] | MTF20 n | MTF20 [lp/mm] |
| --- | --- | --- | --- | --- | --- |
| C1 | 0.2 | 980 | 0.316 (0.294–0.341) | 487 | 2.870 (2.739–2.995) |
| C2 | 0.2 | 980 | 0.339 (0.314–0.369) | 485 | 2.727 (2.584–2.875) |
| C3 | 0.2 | 980 | 0.399 (0.355–0.450) | 484 | 2.451 (2.231–2.667) |
| C4 | 0.2 | 980 | 0.483 (0.410–0.594) | 485 | 2.054 (1.806–2.425) |
| C5 | 0.2 | 980 | 0.496 (0.405–0.602) | 485 | 2.059 (1.780–2.434) |
| C1 | 0.4 | 492 | 0.567 (0.532–0.600) | 245 | 2.861 (2.765–2.937) |
| C2 | 0.4 | 492 | 0.575 (0.534–0.614) | 245 | 2.712 (2.624–2.800) |
| C3 | 0.4 | 492 | 0.619 (0.570–0.676) | 245 | 2.439 (2.309–2.548) |
| C4 | 0.4 | 492 | 0.687 (0.605–0.770) | 245 | 1.970 (1.818–2.148) |
| C5 | 0.4 | 491 | 0.697 (0.608–0.802) | 245 | 1.992 (1.830–2.163) |

Values are medians (interquartile ranges) from QIR 1 reconstructions. SSP n counts slice-wire measurements; MTF20 n counts slices. Counts reflect endpoint-specific availability and exclusion of failed SSP fits.

Variability across slices generally increased with faster configurations. In C5 relative to C1, SSP IQR increased from 0.047 to 0.197 mm and MTF20 IQR increased from 0.256 to 0.654 lp/mm (both P<.001). MTF50 showed the opposite pattern: its IQR decreased by 18%–22% in C4–C5 relative to C1 at 0.2 mm and by 26%–29% at 0.4 mm (Fig S2).

Increasing slice thickness from 0.2 to 0.4 mm increased SSP by a median 0.221 (0.158–0.277) mm across 2459 matched slice-wire measurements (P<.001). Median MTF20 decreased by only 0.008–0.084 lp/mm (0.3%–4.1%) across configurations (Table 2). Corresponding oblique-wire images appear in Figure S1; mean MTF curves and slice-level MTF50 and MTF20 appear in Figure S2.

From C1 to C5, median MTF50 decreased from 2.146 to 1.365 lp/mm at 0.2-mm reconstruction thickness and from 2.123 to 1.360 lp/mm at 0.4 mm (Fig S2). All pairwise comparisons had P<.001 except C4 versus C5 (P=.691 and .127, respectively).

### Effect of QIR Strength

Increasing QIR strength reduced MTF20 variability across slices, with IQR reductions of 40%–57% (P<.001). Median MTF20 generally decreased; the overall difference was nonsignificant in C3 (P=.145). Median SSP increased by only 0.007–0.055 mm from QIR 0 to the highest available strength (1 for C1 and 2 for C2–C5), despite statistical significance (P<.001). SSP variability did not differ significantly across QIR levels (C4, P=.012; others, P≥.056) (Fig S3). QIR therefore mainly affected MTF20 and its variability, with only small changes in SSP.

Median MTF50 also generally decreased with QIR (all P<.001 except C4, P=.853), and its variability decreased in all configurations (P<.001; Fig S3).

### Simulation of Combined Spatial Resolution

The simulated circle became less sharply defined in faster configurations, with blurring in both x and z and greater elongation along z (Fig 2). The unchanged ideal-object outline shows how the same 0.5-mm object can have different apparent shape and contrast despite identical reconstruction settings. Increasing slice thickness to 0.4 mm predominantly increased z-direction blurring, while the configuration-dependent differences along x remained visible.

### Airway Lumen Detectability

At 0.2-mm slice thickness, the 0.4- and 0.6-mm lumens with 0.2-mm walls were detected in C1 but not in C2–C5 (Figs 3, 4). Figure 4 follows these airway sizes across configurations: the opposing walls became less distinct, and the central lumen was no longer discernible in the faster configurations. At 0.4-mm slice thickness, C1 retained the 0.6-mm lumen, while its 0.4-mm lumen was no longer detected. Longitudinal reformations showed reduced wall separation and less continuous lumen depiction in faster configurations, especially at 0.4 mm (Fig 5). Complete maps and lumen detectability boundaries appear in Figure S4.

**Figure 5.**
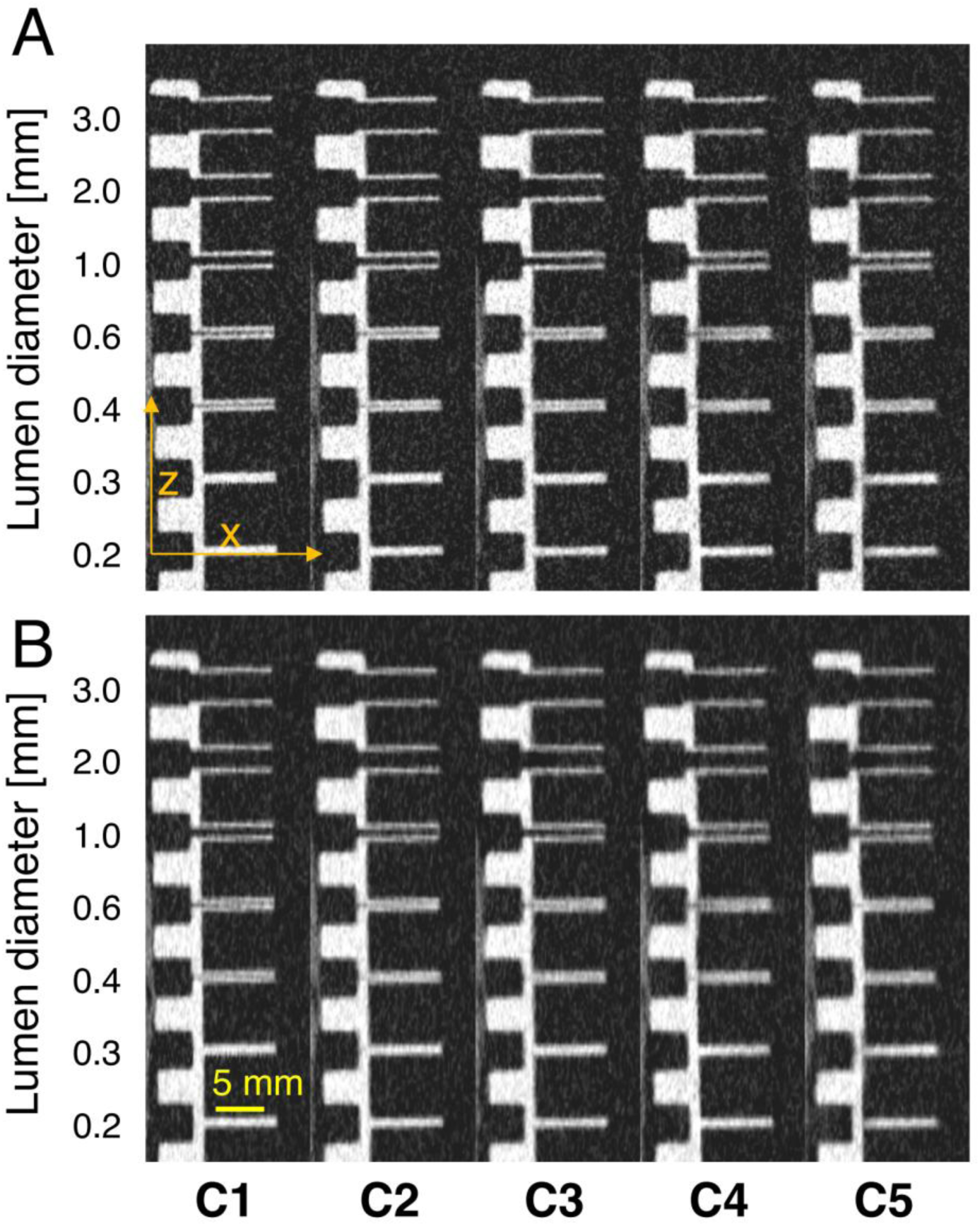
Longitudinal airway depiction across configurations and reconstruction thicknesses. Matched longitudinal reformations through the airway-matrix column with 0.4-mm walls show C1–C5 from left to right. The 0.2-mm reconstructions occupy the upper row and 0.4-mm reconstructions the lower row. Nominal lumen diameters are labeled at left. Smaller airways show reduced separation of opposing walls and less continuous lumen depiction in the faster configurations. Window level/width, −500/1400 HU; scale bar, 5 mm.

For 0.4-mm walls, the 0.4- and 0.6-mm lumens were detected in C1–C3 at 0.2-mm reconstruction thickness. C4 depicted the 0.6-mm lumen but not the 0.4-mm lumen. C5 had an isolated nonmonotonic finding: the 0.4-mm lumen was detected, whereas the 0.6-mm lumen was not. At 0.4-mm reconstruction thickness, both sizes were detected only in C1. A 0.3-mm lumen with 0.5-mm walls was detected only in C1 at both reconstruction thicknesses (Fig S4).

### Venous Stents and Geometric Distortion

Stent-strut separation was clearest in C1 and less distinct in faster configurations (Fig 6A, 6B). The adjacent phantom pipe wall was straight in C1 and geometrically distorted in faster configurations, particularly C3–C5 (Fig 6A, enlarged in Fig 6C). Axial images showed loss of strut definition and contour distortion (Fig S5). Longitudinal views of the additional stents showed similar loss of detail (Figs S6, S7).

**Figure 6.**
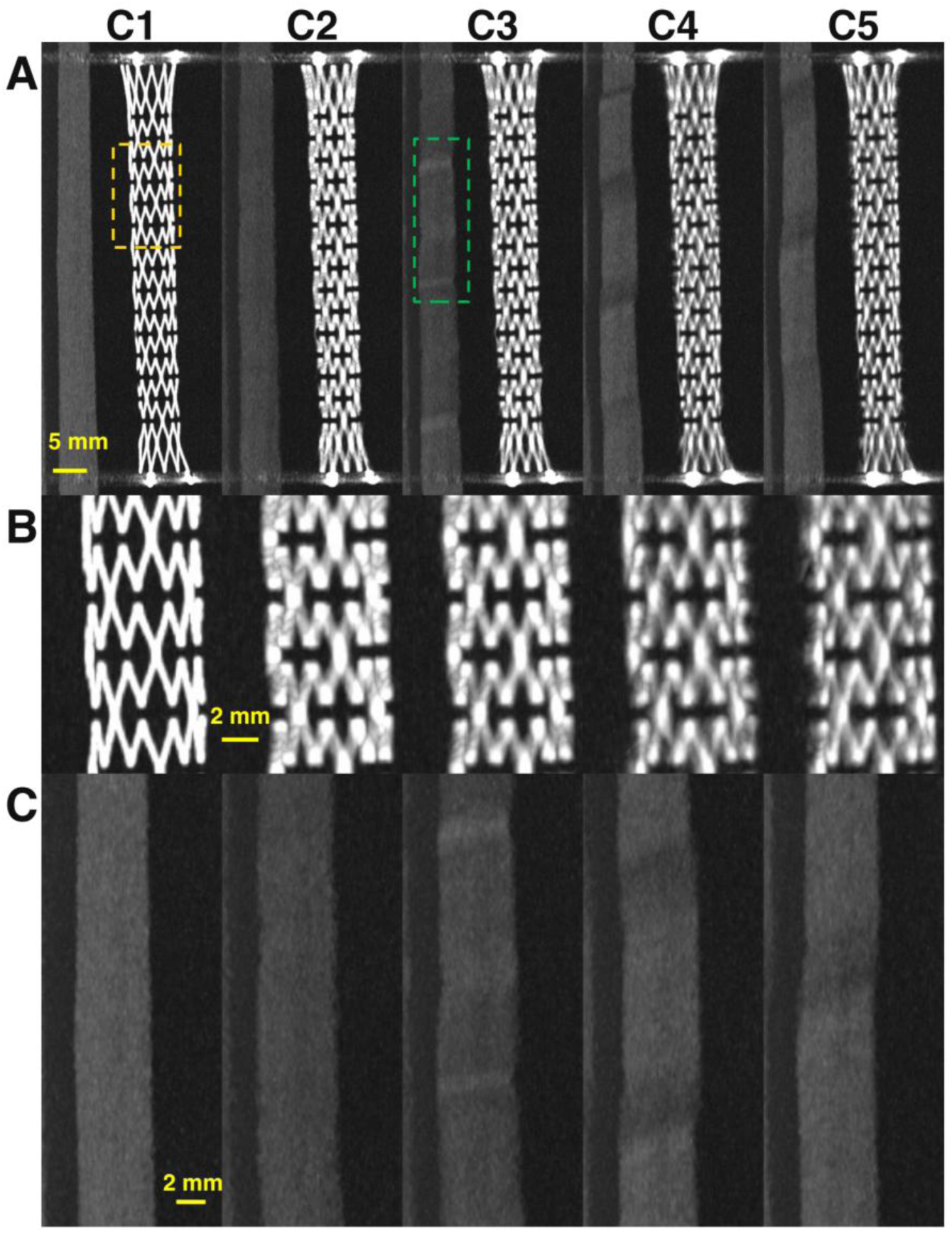
Venous-stent depiction and geometric distortion. (A) Matched 7-mm maximum-intensity projections from 0.2-mm reconstructions show a 10×60-mm venous stent across C1–C5. (B) Corresponding enlargements show stent-strut definition. (C) Corresponding phantom pipe-wall enlargements show repeated contour irregularities, most evident in C3–C5. The yellow dashed box in C1 and green dashed box in C3 identify the stent and pipe-wall source regions, respectively. All rows are aligned by configuration. Window level/width, 1800/4500 HU; scale bars, 5 mm in A and 2 mm in B and C.

## DISCUSSION

Faster ultrahigh-resolution photon-counting CT acquisition configurations were associated with poorer in-plane and through-plane resolution despite identical reconstruction parameters. Variability across slices generally increased, although MTF50 IQR decreased by 18%–29% in C4–C5 relative to C1. The accompanying loss of small-airway and stent detail shows why acquisition configuration matters when choosing a protocol to depict fine structures. A nominal slice thickness or UHR designation alone does not describe the resolution achieved.

The circle simulation helps explain this distinction. A small object is blurred both within the image plane and along the scan direction. In-plane and through-plane resolution therefore jointly determine its apparent shape and contrast. In our simulation, the same circular object appeared more elongated and less sharply defined with faster acquisition configurations. Increasing slice thickness predominantly added through-plane blurring, while in-plane differences persisted. Selecting a thinner slice thus addresses only one component of the loss of detail. The simulation illustrates these combined effects; it does not reproduce the periodic geometric distortion visible in the acquired images.

The airway matrix shows how these differences can affect an imaging task. Small lumens detected with one configuration were no longer detected with another despite unchanged reconstruction settings. The clinically relevant difference is which structures remain distinguishable, particularly near the limit of detection. The loss of stent-strut separation provides a second example of reduced fine-structure depiction. Pulmonary-nodule studies likewise support evaluating the specific imaging task rather than relying on the UHR designation (14).

Configuration-dependent visibility is especially relevant to longitudinal airway assessment. In cystic fibrosis, the RECOVER trial documented improved CT mucus-plugging and bronchial-wall-thickening scores after elexacaftor/tezacaftor/ivacaftor treatment (15). If acquisition or reconstruction settings change between examinations, altered lumen or wall depiction could complicate interpretation of disease progression or treatment response. Consistent technique is therefore important when comparing examinations over time. Our stationary phantom results support this principle; they do not establish clinical detection thresholds.

Changes in helical sampling offer one explanation for the observed configuration differences. At higher pitch, the table advances farther during each rotation, changing the distribution of measured x-ray beams and the interpolation needed to form an image. A second x-ray source in dual-source scanners supplies additional measurements, but does not eliminate the helical artifacts that can occur at high pitch (7, 16). Prior studies have demonstrated pitch-dependent geometric distortion and periodic surface irregularities in stationary phantoms (17, 18). The distortion of the phantom pipe wall and the greater variability across slices are consistent with a contribution from helical sampling. Because the clinical configuration parameters changed together, these observations do not isolate the effect of pitch.

Focal-spot size provides a complementary explanation. At comparable dose, higher pitch or faster rotation requires greater instantaneous tube current, which can enlarge the focal spot and reduce spatial resolution (6). Focal-spot class increased from XS to S and M across the tested configurations. For dual-source acquisition, current per source is the relevant quantity. These changes can affect both in-plane and through-plane resolution. We tested 120 kVp; at matched dose and otherwise fixed acquisition settings, a higher voltage may reduce the required current and limit focal-spot blooming, although blooming can persist at higher currents (6).

The same consideration applies when choosing tin filtration for dose reduction. Tin removes low-energy photons and has enabled low-dose noncontrast PCCT (19, 20). Its dose advantages should be considered alongside possible resolution changes (21). At matched dose and otherwise fixed settings, tin filtration requires higher current, potentially increasing focal-spot blooming. We did not test tin filtration, so this is a protocol consideration rather than a finding of the present study.

Reconstruction strength offered a different tradeoff. Increasing QIR reduced variability in in-plane resolution across slices, while in-plane resolution generally decreased. The changes in through-plane resolution were small, and its variability did not differ significantly across QIR levels at the selected threshold. QIR therefore mainly affected in-plane resolution and its variability, with a limited effect on through-plane resolution. It did not recover the larger differences associated with acquisition configuration. Prior UHR lung PCCT work similarly found noise reduction with QIR and a small reduction in measured resolution (22). We did not characterize image noise across QIR levels; our results describe the resolution component of that tradeoff.

Wider active UHR detector coverage could reduce the need for high pitch or short rotation time to complete a scan within a breath-hold. Coverage of 40–80 mm could permit lower pitch or slower rotation at a chosen acquisition time, provided detector sampling and focal-spot performance are maintained (3). This potential benefit should be distinguished from the two pitch-3.2 configurations tested here, which showed similarly degraded resolution and a small acquisition-time difference. The narrower Flash UHR mode also permits additional standard-resolution spectral reconstructions (23). Coverage, spectral capability, and the resolution required for the task therefore belong in the same protocol decision.

These helical-acquisition findings do not directly apply to cardiac UHR imaging on this scanner, which uses ECG-triggered axial step-and-shoot or low-pitch helical acquisitions. Focal-spot size remains relevant, but the coverage constraint creates a different problem: multiple heartbeats are needed to image the heart, introducing sensitivity to beat-to-beat variability and changes in contrast enhancement. Reconstruction tools such as ZeeFree can reduce misalignment between adjacent image stacks (24). Contrast differences between acquisitions require separate consideration.

This study has limitations. We tested one scanner and one quantitative kernel in stationary phantoms. Cardiac and respiratory motion may alter the balance between acquisition time and spatial resolution. All objects were within 10 cm of isocenter; peripheral resolution was not evaluated and can be lower (25). The oblique airway matrix reflected combined in-plane and through-plane resolution at one position and orientation. We used an objective profile-based detection criterion without a reader study; agreement with radiologist detection remains untested. Characterizing nonlinear reconstruction additionally requires accounting for object contrast and noise (26). Measurements characterized within-scan variability, and stent assessment was qualitative.

In conclusion, faster ultrahigh-resolution photon-counting CT configurations were associated with poorer in-plane and through-plane resolution and greater through-plane variability across slices. The accompanying loss of small-airway and stent detail supports task-specific protocol selection and consistent acquisition and reconstruction settings for longitudinal imaging. Nominal slice thickness and the ultrahigh-resolution label alone do not describe the spatial resolution achieved and should be used and reported with caution.

## Abbreviations

IQR: interquartile range
MTF: modulation transfer function
PCCT: photon-counting detector CT
QIR: quantum iterative reconstruction
SSP: slice sensitivity profile
UHR: ultrahigh resolution.

## Data Availability

Data generated or analyzed during this study are available from the corresponding author upon reasonable request.

