## Supplementary Material for "Acquisition Speed versus Spatial Resolution in Ultra-High-Resolution Photon-Counting CT: Phantom Study to Guide Protocol Development"

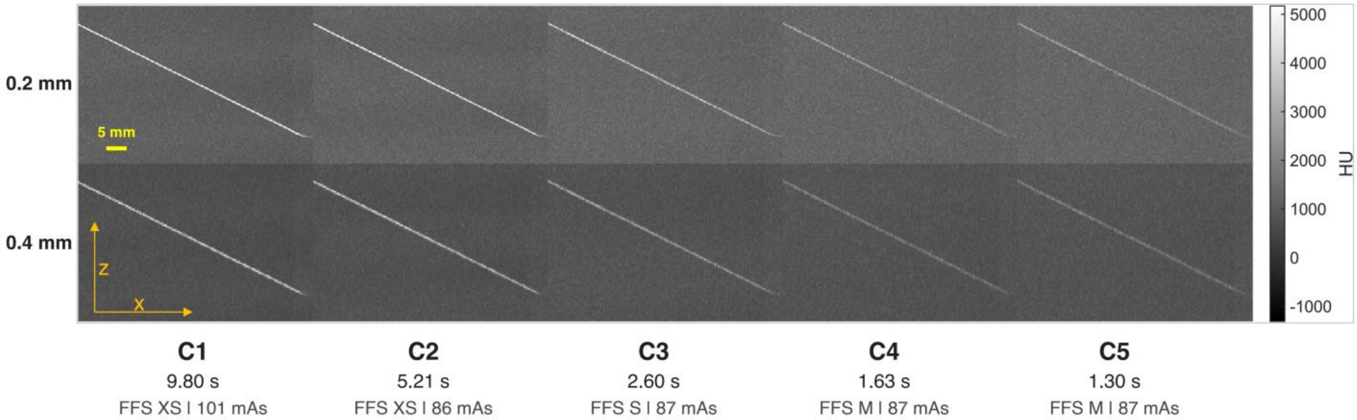


Figure S1. Appearance of an oblique wire across UHR acquisition configurations. Coronal x-z maximum-intensity projections through the same oblique wire are shown for 0.2-mm (top) and 0.4-mm (bottom) reconstructions using QIR 1. C1–C5 are ordered from longest to shortest calculated acquisition time for 400-mm coverage. Labels report calculated time, focal-spot class, and effective exposure for the wire acquisitions. All panels use identical display limits. The wire changes appearance with acquisition configuration and reconstruction thickness. These projections were not used to calculate SSP or MTF. Yellow scale bar, 5 mm. FFS = focal-spot size; MTF = modulation transfer function; QIR = quantum iterative reconstruction; SSP = slice sensitivity profile; UHR = ultra-high resolution.

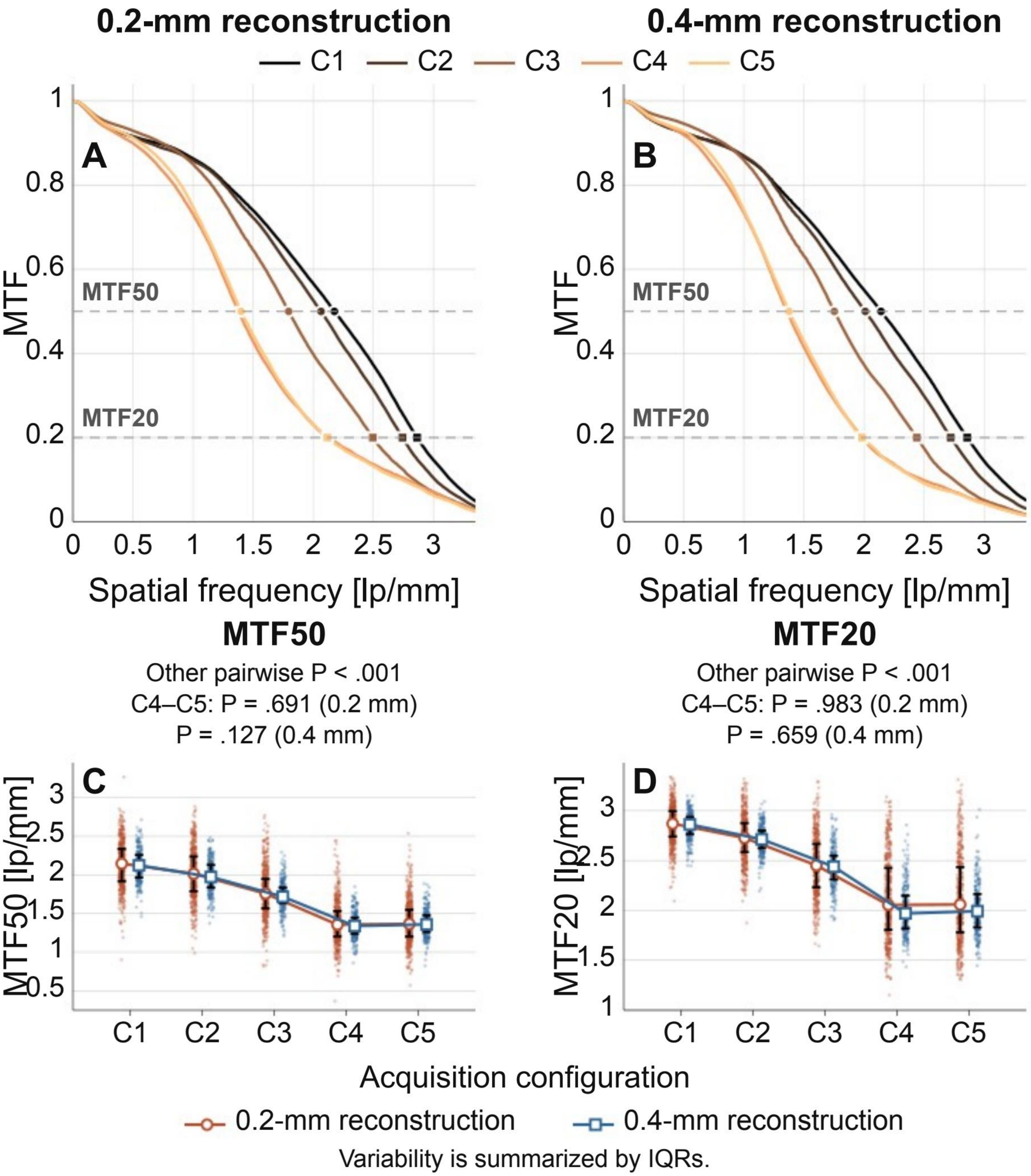


Figure S2. Mean MTF curves and variability across slices. Mean MTF curves are shown for 0.2-mm (A) and 0.4-mm (B) reconstructions across C1–C5 using QIR 1. Circles and squares identify MTF50 and MTF20 read from the mean curves. Slice-level MTF50 (C) and MTF20 (D) are shown for both thicknesses. Points show individual measurements; colored lines and markers show medians, and black error bars show IQRs. Labels report Holm-adjusted configuration comparisons. Mean-curve thresholds and medians of slice-level thresholds are distinct summaries. IQR = interquartile range; MTF = modulation transfer function; QIR = quantum iterative reconstruction.

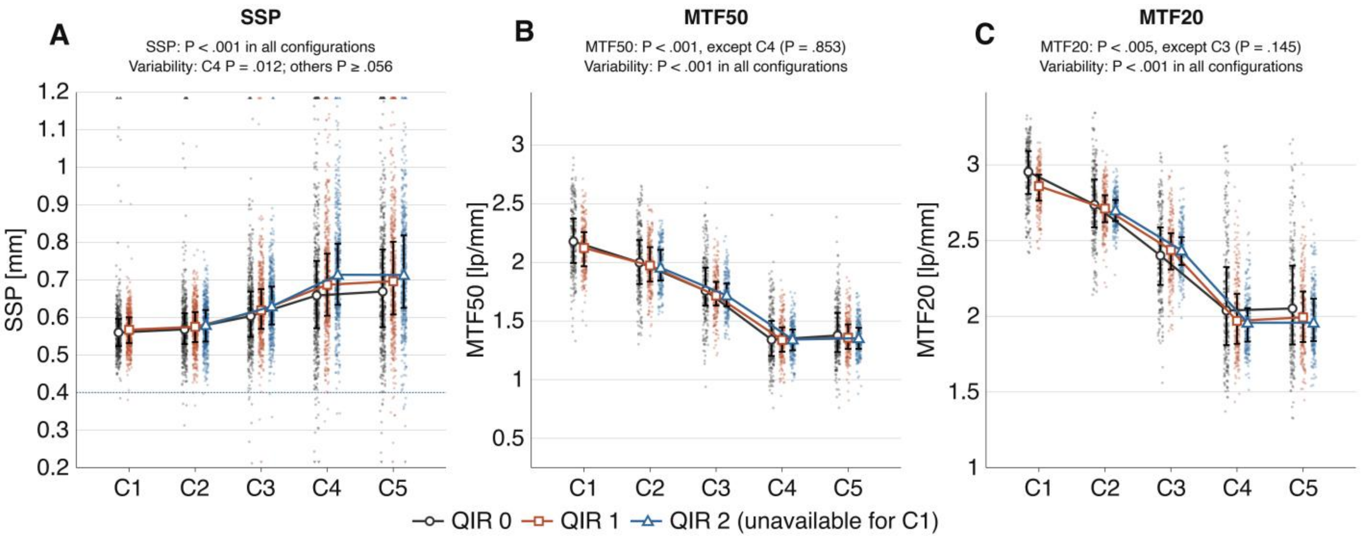


Figure S3. Effect of QIR strength on spatial resolution and variability across slices. Slice-level SSP (A), MTF50 (B), and MTF20 (C) are shown for 0.4-mm reconstructions across C1–C5. Points show individual measurements; colored lines and markers show medians, and black error bars show IQRs. Triangles at axis limits identify observations outside the displayed range, retained in analysis. The dotted line in A indicates nominal reconstruction thickness. QIR 2 was unavailable for C1. Labels report overall Holm-adjusted P values for differences across QIR levels and variability across slices. SSP increases were very small despite statistical significance. SSP variability did not differ significantly across QIR strengths at P<.01, including C4 (P=.012). IQR = interquartile range; MTF = modulation transfer function; QIR = quantum iterative reconstruction; SSP = slice sensitivity profile.

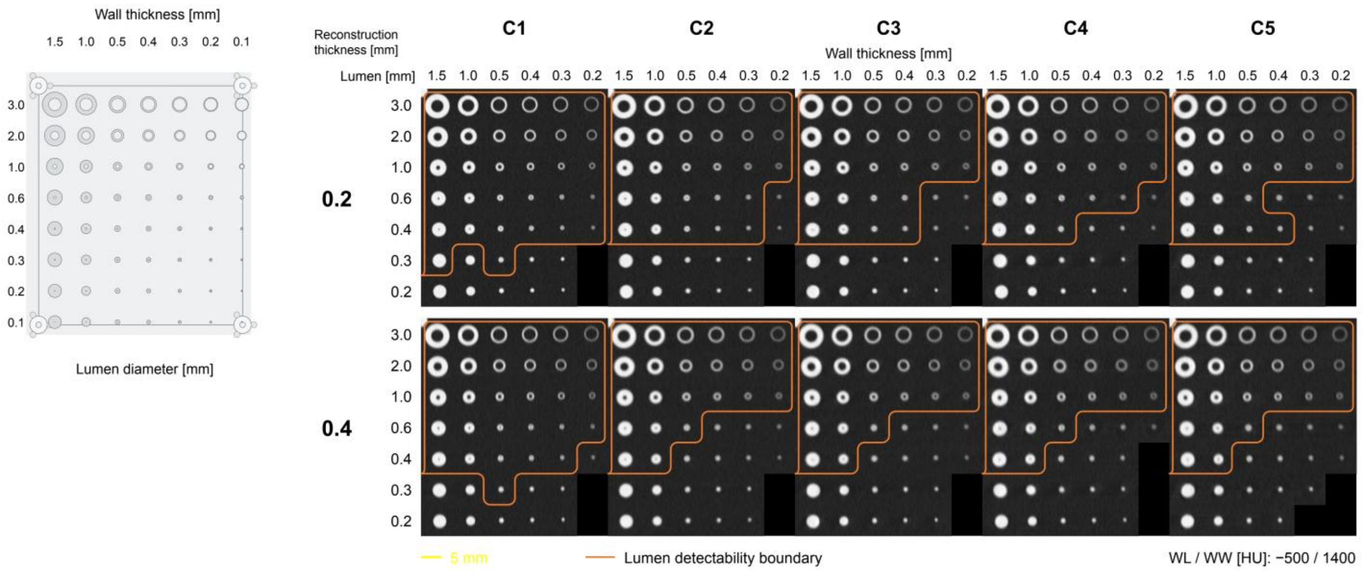


Figure S4. Airway matrix and lumen detectability boundaries at two reconstruction thicknesses. The full design schematic is shown at left. Matched 5-mm oblique mean-intensity reformations show C1–C5 from left to right, with 0.2-mm reconstructions above 0.4-mm reconstructions. Orange closed contours delineate regions containing lumens that met the detection criterion at the sampled positions. Curves are visual guides, retain isolated nonmonotonic findings, and do not represent fitted continuous thresholds. Damaged or unevaluable positions are omitted from CT panels and retained in the schematic. Images share magnification and window level/width of −500/1400 HU. Yellow scale bar, 5 mm.

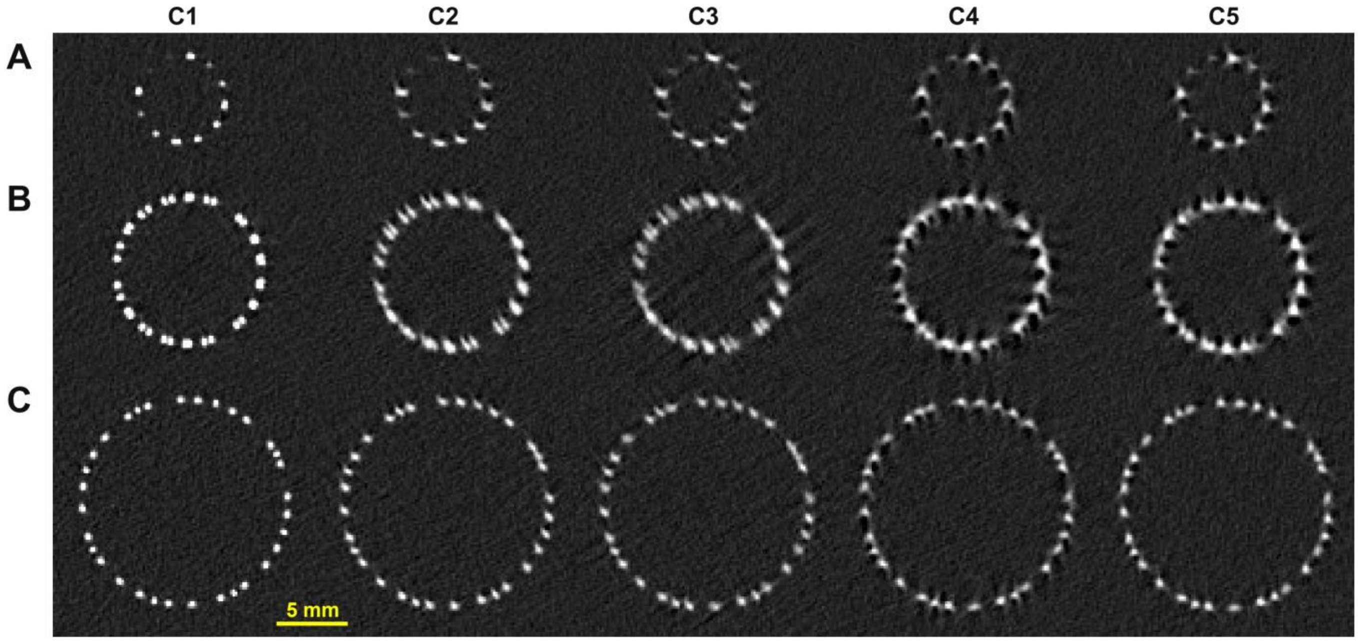


0.2-mm reconstruction | WL / WW [HU]: 2000 / 6000

Figure S5. Axial stent cross-sections. Axial reformations from 0.2-mm reconstructions show three stents, ordered by increasing apparent cross-sectional size from A to C. Columns show C1–C5. Images were sampled at a common axial plane by interpolation between neighboring source slices. All panels share magnification and window level/width of 2000/6000 HU. Yellow scale bar, 5 mm.

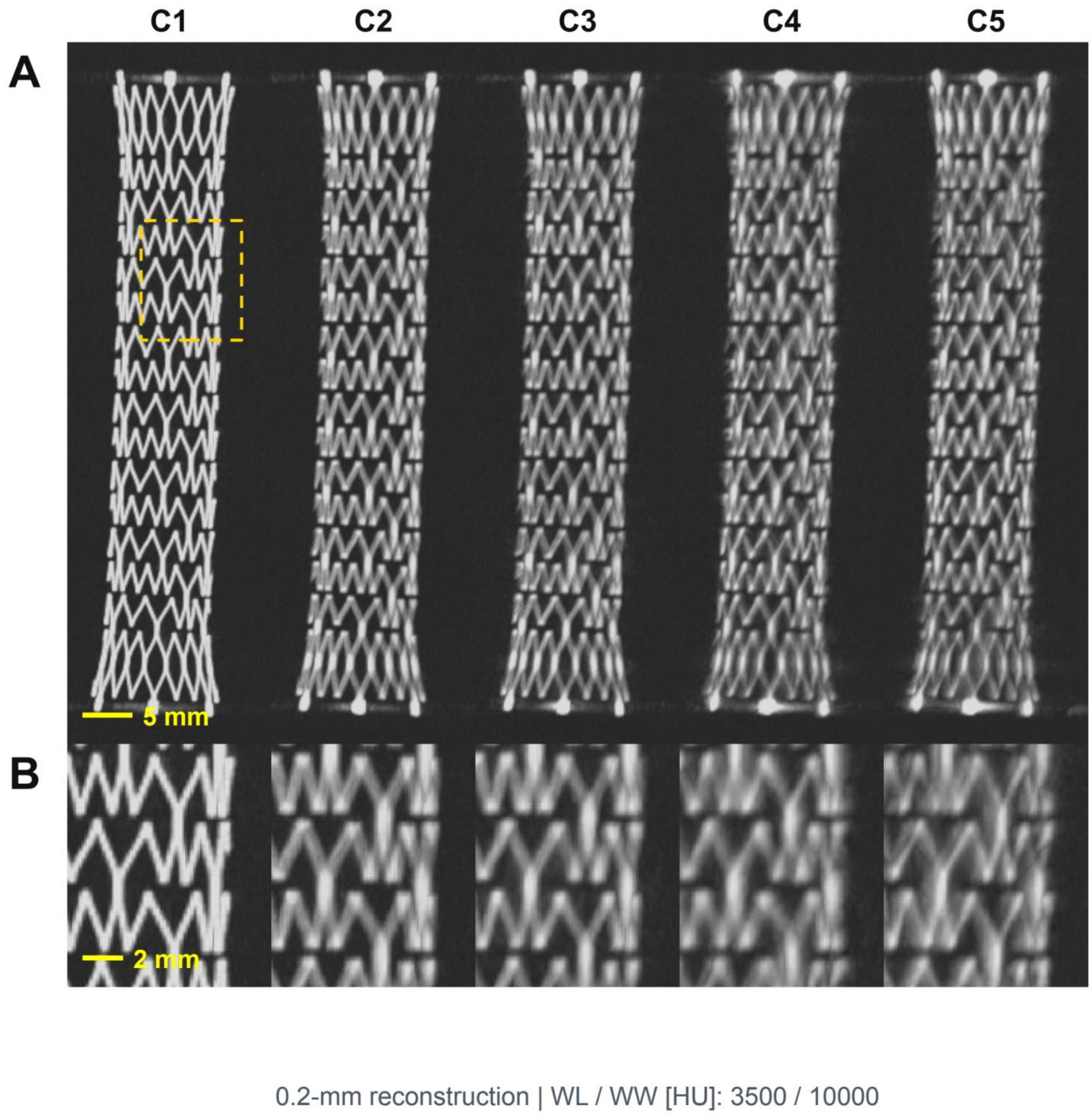


Figure S6. Additional venous stent. Longitudinal maximum-intensity projections from 0.2-mm reconstructions show the stent overview (A) and corresponding enlargements (B) across C1–C5. The yellow dashed box in C1 identifies the region enlarged in B; corresponding regions are enlarged for C2–C5. Images share magnification within each row and window level/width of 3500/10000 HU. Yellow scale bars, 5 mm in A and 2 mm in B.

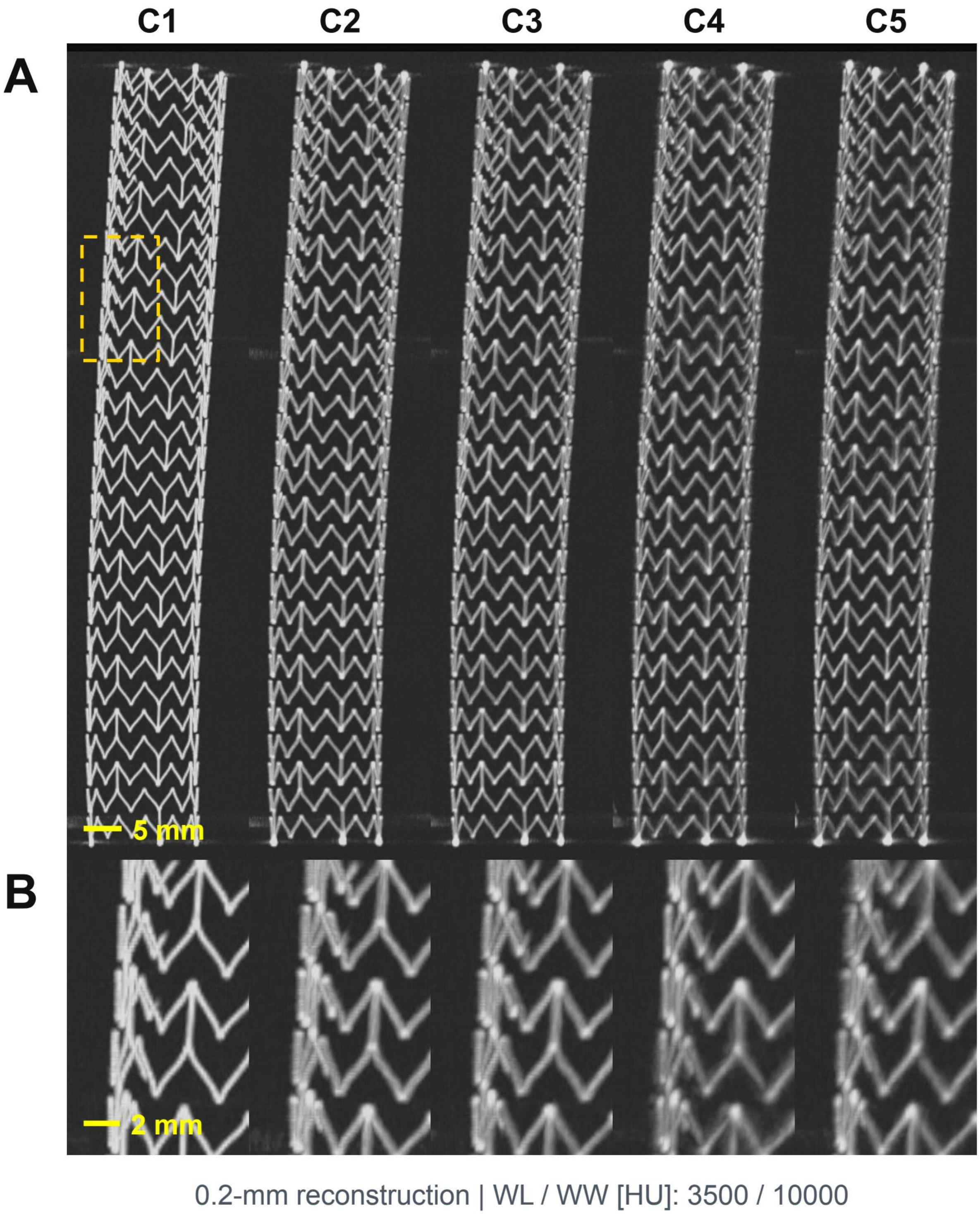


Figure S7. Largest venous stent. Longitudinal maximum-intensity projections from 0.2-mm reconstructions show the stent overview (A) and corresponding enlargements (B) across C1–C5. The yellow dashed box in C1 identifies the region enlarged in B; corresponding regions are enlarged for C2–C5. Images share magnification within each row and window level/width of 3500/10000 HU. Yellow scale bars, 5 mm in A and 2 mm in B.
